# Computer skills of health workers in rural Ghana and their use of social media: Implications for increased use of information technology in service delivery

**DOI:** 10.1101/2021.10.16.21265098

**Authors:** Sylvia K Ofori, Emmanuel A Akowuah, Doyinsola Babatunde, Logan Cowan, Frank Baiden

## Abstract

**Background:** Information and Communication Technologies (ICTs) have improved the delivery of healthcare worldwide especially in disease control, patient management, and health data analysis. However, access to infrastructure like computers and knowledge of ICT is a major problem to various digital health initiatives among health professionals.

**Objective:** The aim of the present study was, to assess healthcare workers’ basic computer skills and use of social media and identify specific deficiencies that require training to help them function effectively.

**Methods:** We conducted a cross-sectional study among 51 healthcare workers in seven health centers in a region in Ghana over three months using a self-administered pre-tested questionnaire.

**Results:** Half of the participants had adequate basic computer skills, of which 60.87% were women (P=0.0205). Only 15.69%, who were all males, of participants used computer software at least once a month. Most participants also used Facebook and WhatsApp.

**Conclusion:** Health professionals in the health centers need to be trained on basic computer skills and, information technology should be incorporated into the health systems for efficient health delivery.

## Introduction

Information and Communication Technologies (ICTs) are devices and technologies that access, process, store, manipulate and transmit information electronically in digital form.^1, 2^ Factors that influence the use of ICT include knowledge, attitude, and access. The advent of technology has improved the delivery of healthcare worldwide especially in disease control, patient management, and health data analysis.^3^ Information technology has made it easier for the dissemination of health information worldwide, clinical data management, and strengthening of health systems, especially in developing countries. However, access to infrastructure like computers and knowledge of ICT is a major problem to various digital health initiatives among health professionals.^4^ Developing countries also face limited and expensive broadband access especially in Africa where about 46 countries share only 25% of internet hosts.^5^ Only 28% of health facilities in Ethiopia use computers.^6^ It has been reported that even though most health facilities in Ghana have computing equipment, multimedia device, and internet systems, these infrastructures have not been fully integrated to support health delivery.^7^ A study to assess health workers’ knowledge of computer applications in rural health facilities in Ghana and Tanzania reported that only 40% had ever used computers with the majority of them (80%) being computer illiterates or beginners.^8^ However, in their study, the frequency of computer use or the need to use a computer in their work was not explored. The majority of nurse managers (85%) in a hospital in Kenya were also found to have received no computer training in their basic nursing training with almost all of them desiring to be trained in computer skills.^9^ However, their study was conducted in a single hospital and hence was limited in generalizability. Health workers in primary health centers in Ethiopia were found to have lower knowledge (3.4%) and use of computers (18.4%) indicating a disparity depending on the level of healthcare.^4^

Information and Communication Technologies can also play a vital role in health service delivery to children. One way to achieve this is by enabling key healthcare workers to perform duties by improving access to information and improving accountability. Basic computer skills can help health workers efficiently collect data, perform basic analysis, store and disseminate health-related information.^4^ This will help assess trends of diseases and, also routinely assess the performance of child health indicators including neonatal mortality rate, immunization uptake rates, and incidence of infections over a period. The use of social media by health workers can help them to access and share relevant information, engage with the public and patients, develop a professional network and increase awareness of health-related discoveries.^10, 11^ Thus, these skills can enable health centers in rural areas to perform continuous quality improvement to improve the health of children.

Most of the studies assessing healthcare workers’ computer skills and use of social media have focused on knowledge, use, and attitude. ^3, 12, 13^ No study was found to assess the skills of health workers in specific computer skills or find the association between social media use and computer skills. Also, only a few studies were found to have conducted their research on multiple sites. Hence, our study was conducted to address these gaps in the literature.

The aim of the present study was, therefore, to assess health workers’ basic computer skills and use of social media and identify specific deficiencies that require training to help them effectively function as above. We hypothesized that there is an association between profession and years working at a health facility, and basic computer skills or frequency of using computer programs.

### Materials and methods Ethical Considerations

The study was approved by the Institutional Ethics Committee of Kintampo Health Research Centre and the Ethics Review Committee of Ghana Health Service. Approvals were obtained from the District and Municipal Health Management Teams and the health center managers. Signed individual informed Consent was obtained from each participant after they reviewed information sheets provided by research participants. Information sheets included details on study objectives, procedures, and possible outcomes of the study. Participants had the right to withdraw from the study at any time during data collection without any consequences. Questionnaires retrieved from the study and softcopies of entered data are stored in strict confidence.

### Study Design and population

A cross-sectional study design was used to assesses the use of social media, basic computer skills, and frequency of using computer programs among healthcare workers in health facilities in the Brong Ahafo Region of Ghana over three months. All healthcare workers, a total of 51 in seven health centers were included in the study. This number excluded those who were on annual or sick leave. Health workers were defined as people with a form of clinical training who worked in the health centers including community health workers, medical assistants, and nurse-midwives.

### Study Site

Ghana is a middle-income country in Sub-Saharan Africa with a population of about 28 million, with ten administrative regions. Brong-Ahafo is one of the ten regions that lie in the middle belt. Health centers in this region of Ghana served as the sites for this study. Health centers are the first level of institutional care that serves most of the population especially those in the rural areas. Healthcare workers in these centers include medical assistants, nurse midwives, and community health nurses. They provide services to the community including immunization of children, family planning, first aid, and management of diseases like malaria, upper respiratory tract infections, and diarrhea. Health centers located in six districts including Kintampo North, Kintampo South, Nkoranza North, Nkoranza South, Techiman, and Tain were involved in this study.

### Data collection and analysis

The data for this study was collected in 2014 during an implementational study to support the implementation of test-based management of malaria in under-five children in the Brong-Ahafo Region of Ghana using a quality improvement approach. A pre-tested, self-administered questionnaire was used to collect data on sociodemographic characteristics, ownership of computer and social media accounts, knowledge of statistical software, and use of Microsoft programs in daily life. The questionnaire was available in the English language and collected over two months. Names of participants were not required on the questionnaire to increase participation and ensure anonymity. The data collection took place at the workplace of participants at their own convenient time. Data collection was facilitated by research assistants under the supervision of the principal investigator. To increase the response rate, incentives in the form of hospital supplies were provided by the researchers.

#### Exposure

The exposures of interest in the study were the profession of participants and years of working in the facility. Health professionals or healthcare workers were defined as persons with a form of medical training that provides clinical service in these facilities. The professions include community health officers, nurses, and medical health assistants. Professions that were not common were grouped as others and included interns, nursing assistants, and medical counter assistants. Years worked at the facility were categorized as less than five years, between 6 and 10 years, and more than 11 years. Exposure information was collected with the self-administered questionnaire.

#### Outcome

The outcomes included ownership of computer/laptop, use of social media, basic computer skills including use of Microsoft Office and knowledge of statistical software, and frequency of using computer programs. Computer skills were defined as the use of Microsoft Word, Microsoft Excel, Microsoft PowerPoint, and statistical software in their work activities. Social media use was defined as having an account on Facebook, WhatsApp, and an email address. Frequent users were defined as those who used the software more than once in a month and non-frequent users were those who never used or never heard of the software. Software assessed included Microsoft Word, Excel, PowerPoint, Stata, SPSS, and Epi-info. Participants were also assessed on basic computer skills and classified as having adequate computer skills or not. People were categorized into adequate skills if they used Microsoft Excel for calculations and graphs, and Microsoft PowerPoints for presentation. To be classified as having adequate computer skills, participants must have answered positively to using all the above.

Data was entered using Stata then exported to Statistical Analysis Software (SAS), version 9.1 of the SAS System for Windows for analysis. Descriptive analysis using frequency tables and cross-tabulations were used to summarize the data collected. Percentages were used to represent findings from continuous data. Bivariate analysis was used to explore the relationship between the exposures and outcomes. Fisher’s exact statistics were computed to determine association with the use of social media and other factors (i.e., age, and gender). The chi2 square test was also used to determine the association between basic computer skills and other variables.

## Results

### Sociodemographic Characteristics of study participants (Table 1)

Fifty-one healthcare workers participated in the study, including 21 (41.18%) nurses, 8 (15.69%) community health officers, 2 (3.98%) medical assistants, and 20 auxiliary workers. The age range of participants was between 23 and 62 years old and more than half (29 (56.87%) of the participants were males. The majority of participants (72.55%) had worked for less than five years in their present facilities with only a third of them having a bachelor’s degree or diploma. Most of the participants were from Nkoranza, North, Kintampo North, Techiman South, and Tain Districts.

**Table 1:** Characteristics of the Population.

| **Attribute** | **N (%)** |
| --- | --- |
| **Gender** |  |
| Male | 29 (56.86) |
| Female | 22 (43.14) |
| **Age (years)** |  |
| <34 | 36 (70.59) |
| 35-49 | 6 (11.76) |
| 50-65 | 9 (17.65) |
| **Education level** |  |
| University | 5 (9.8) |
| Diploma | 11 (21.57) |
| Certificate | 22 (43.14) |
| Other (advanced diploma, high school, O'level) | 13 (25.49) |
| **Profession** |  |
| Medical assistant | 2 (3.92) |
| Nurse/midwife | 21 (41.18) |
| Community health officer | 8 (15.69) |
| Other | 20 (39.22) |
| **District** |  |
| Kintampo N | 10 (19.61) |
| Techiman N | 6 (11.76) |
| Tain | 9 (17.65) |
| Nkoranza S | 8 (15.69) |
| Techiman S | 9 (17.65) |
| Nkoranza N | 9 (17.65) |
| **Years worked at the facility** |  |
| 0-5 | 37 (72.55%) |
| 6- 10 | 7 (13.73%) |
| 11 years or more | 7 (13.73%) |

### Computer Ownership, Use of Microsoft Software and Social Media (Table 2)

About half of the respondents (n=25) had personal laptops with only 19 (37.25%) having access to desktops at the office. The majority of participants had personal email addresses 32 (62.75%), Facebook (31(60.78%), and WhatsApp (36 (70.595). Less than half of participants had skills in using Microsoft Excel as calculators 18 (35.29%) or for graphs 20 (39.22%), and PowerPoint for presentations (N= 25 (49.02%).

**Table 2.**
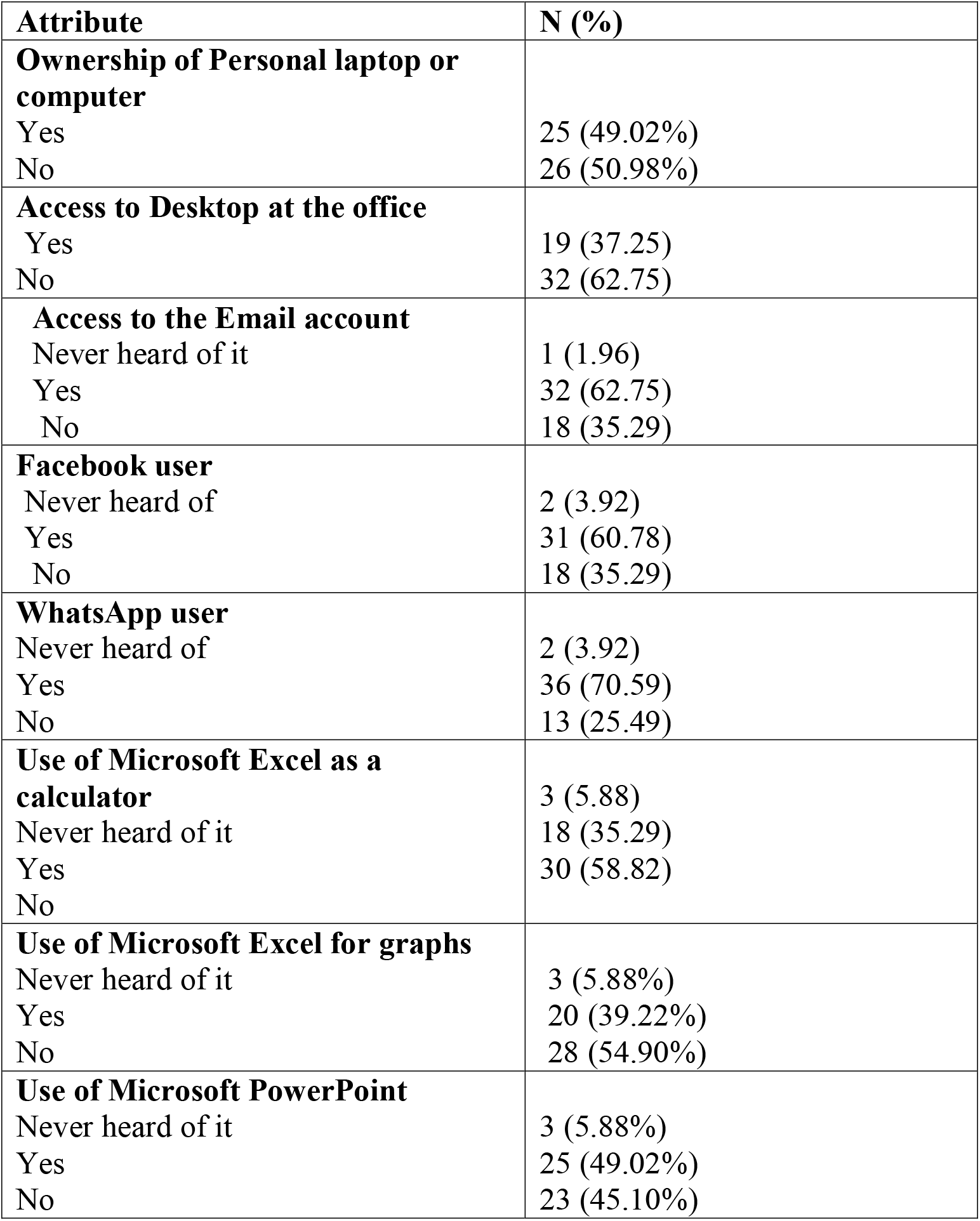
Computer Ownership, Use of Microsoft Software and Social Media.

| **Attribute** | **N (%)** |
| --- | --- |
| **Ownership of Personal laptop or computer** |  |
| Yes | 25 (49.02%) |
| No | 26 (50.98%) |
| **Access to Desktop at the office** |  |
| Yes | 19 (37.25) |
| No | 32 (62.75) |
| **Access to the Email account** |  |
| Never heard of it | 1 (1.96) |
| Yes | 32 (62.75) |
| No | 18 (35.29) |
| **Facebook user** |  |
| Never heard of | 2 (3.92) |
| Yes | 31 (60.78) |
| No | 18 (35.29) |
| **WhatsApp user** |  |
| Never heard of | 2 (3.92) |
| Yes | 36 (70.59) |
| No | 13 (25.49) |
| **Use of Microsoft Excel as a calculator** |  |
| Never heard of it | 3 (5.88) |
| Yes | 18 (35.29) |
| No | 30 (58.82) |
| **Use of Microsoft Excel for graphs** |  |
| Never heard of it | 3 (5.88%) |
| Yes | 20 (39.22%) |
| No | 28 (54.90%) |
| **Use of Microsoft PowerPoint** |  |
| Never heard of it | 3 (5.88%) |
| Yes | 25 (49.02%) |
| No | 23 (45.10%) |

### Computer skills of respondents and social media (Table 3)

Generally, about half of the participants had adequate computer skills. More females had adequate computer skills compared to men (p=0.0205) and the majority of participants in the adequate computer skills category were less than 34 years old (N= 14 (60.87%) followed by those between 35 and 49 years (5(21.74%), but the difference between groups was not significant. The majority of those with inadequate skills also were below 34 years. More nurses had adequate computer skills 10 (43.48%) compared to the other professions and almost the same number of nurses had inadequate computer skills. The majority of people with adequate computer skills had spent less than five years at the facility. People with personal laptops and access to desktops at the office had adequate computer skills.

**Table 3:** Computer skills of respondents and social media.

| **Attribute** | **Adequate**<br>**Computer skills**<br>**=N (%)** | **Inadequate skills**<br>**= N (%)** | **P. value** |
| --- | --- | --- | --- |
| **Gender**<br>Male<br>Female | 9 (39.13)<br>14 (60.87) | 20 (71.43)<br>8 (28.57) | **0.0205** |
| **Age**<br><34<br>35-49<br>50-65 | 14 (60.87)<br>5 (21.74)<br>4 (17.39) | 22 (78.57)<br>1 (3.57)<br>5 (17.86) | 0.1423 |
| **Profession**<br>Medical assistant<br>Nurse/midwife<br>Community health officer<br>Other | 1 (4.35)<br>10 (43.48)<br>2 (8.70)<br>10 (43.48) | 1 (3.57)<br>11 (39.29)<br>6 (21.43)<br>10 (35.71) | 0.6968 |
| **Education**<br>University<br>Diploma<br>Certificate<br>Other (advanced diploma,<br>high school, O'level) | -<br>4 (17.39%)<br>12 (52.17%)<br>7 (30.4%) | 5 (17.86%)<br>7 (25.00%)<br>10 (35.71%)<br>6 (21.43%) | 0.4681 |
| **Years spent at the facility**<br>0-5<br>6- 10<br>11 years or more | 17 (73.91%)<br>3 (13.04%)<br>3 (13.04%) | 20 (71.43%)<br>4 (14.29%)<br>4 (14.29%) | 1.00 |
| **Ownership of Personal**<br>**laptop or computer**<br>Yes<br>No | 9 (39.13%)<br>14 (60.87%) | 16 (57.14%)<br>12 (42.86%) | 0.26 |
| **Access to Desktop at the**<br>**office**<br>Yes<br>No | 10 (43.48%)<br>13 (56.52%) | 9 (32.14%)<br>19 (67.86%) | 0.29 |

### Frequency of use of computer programs (Table 4)

The frequency of using computer software was low, as only 15.69% (n=8) of participants used the software at least once a month. All frequent users of computer programs were males even though almost half of the males were classified as non-frequent users (p. value= 0.0072). The majority of infrequent users were nurses and community health workers. Only those below 34 years old and who had spent less than five years in the facility were frequent users of computer programs. People with certificate qualifications formed a major part of the infrequent users. People with laptops and desktops formed the majority of routine users even though the association was not significant.

**Table 4:** Frequency of use of computer programs.

| **Attribute** | **Routine users** | **Non-routine users** | **P. value** |
| --- | --- | --- | --- |
| **Gender** |  |  |  |
| Male | 8 (100) | 21 (48.84) | **0.0072** |
| Female | -- | 22 (51.16) |  |
| **Age (yrs)** |  |  | 0.2206 |
| <34 | 8 (100) | 28 (65.12) |  |
| 35-49 | --- | 6 (13.95) |  |
| 50-65 | ---- | 9 (20.93) |  |
| **Profession** |  |  | 0.3758 |
| Medical assistant | 1 (12.50) | 1 (2.33) |  |
| Nurse/midwife | 3 (37.50) | 18 (41.86) |  |
| Community health officer | 2 (25.00) | 6 (13.95) |  |
| Other | 2 (25.00) | 18 (41.86) |  |
| **Education level** |  |  | 0.4681 |
| University | 2 (25.00) | 3 (6.98) |  |
| Diploma | 1 (12.50) | 10 (23.26%) |  |
| Certificate | 3 (37.50%) | 19 (44.19) |  |
| Other (advanced diploma, high school, O'level) | 2 (25.00) | 11 (25.58) |  |
| **Years spent at the facility** |  |  | 0.3935 |
| 0-5 | 6 (75%) | 31 (72.09%) |  |
| 6- 10 | 2 (25%) | 5 (11.63%) |  |
| 11 years or more | - | 7 (16.28%) |  |
| **Ownership of Personal laptop or computer** |  |  | 0.14 |
| Yes |  |  |  |
| No | 6 (75%)<br>2 (25%) | 19 (44.19%)<br>24 (55.81%) |  |
| **Access to Desktop at office** |  |  | 0.13 |
| Yes |  |  |  |
| No | 5 (62.50%)<br>3 (37.50%) | 14 (32.56%)<br>29 (67.44%) |  |

## Discussion

The findings of the study show that the frequency of using a computer and social media, and computer skills were generally low for healthcare workers in primary health centers in the Brong Ahafo Region of Ghana. The results were lower than in other studies.^14, 15^ However, the findings on computer skills were higher than that reported in a study among healthcare workers in Addis Ababa Hospitals in Ethiopia^16^ and staff in a Nigerian University teaching hospital.^3^ The differences may be due to socio-economic differences among study participants.

The results of the association between ownership of laptops and frequency of use or knowledge on the computer are consistent with other studies.^16^ The finding that participants with adequate computer skills were likely to be females is inconsistent with other studies^14, 17^ although the association was not significant in our study. This may be because more females owned laptops and had access to desktops in their offices. We also found that younger healthcare workers and those who had spent less than five years had adequate skills and this result agreed with previous studies. ^4, 14, 18^ This finding suggests that training programs should be organized for older professionals. Level of education was not significantly associated with computer skills and frequent use of the computer which was consistent with studies in India.^14^

Unlike other studies that reported a significant association between knowledge and sex, profession, age, ownership of laptops, our study only found gender to be significant.^19^ In our study, the use of electronic mail and the internet was high which is higher than that reported in the study by Adeleke et al.^19^ In their study about 51% of participants used electronic mail compared to over 60% in our study. However, a higher proportion of them had better computer knowledge and skills compared to our study.^19^ Another study reported that almost all the participants had access to the internet (98%) and the use of emails was similar to our study.^20^

Our study also shows that the use of Facebook and WhatsApp was high and this was consistent with a study in Saudi Arabia that reported that the majority of participants used WhatsApp, however the use of Facebook was low compared to our study.^21^ A study reported that Facebook was the third most common social media used by physicians, and the participants reported benefits including access to accurate information and interaction with providers outside the medical facilities.^22^ Other advantages of using social media include peer support, increased interaction with others, and public health surveillance.^2^ Some disadvantages of the use of social media are jeopardizing professional image when questionable content is posted and productivity loss due to spending working hours on these sites.^23^ Most of the studies on the use of social media among health professionals have focused on physicians and our study provides a landscape of use among other health professionals in primary healthcare facilities.^24^ Our results are comparable to other studies, an example is a study to assess the use of Facebook among pharmacist preceptors who reported a prevalence of about 60%.^25^ We did not evaluate the frequency of using social media in a day or the purpose for which it was used. But other studies have reported that Facebook was used mainly for chatting, uploading pictures, and interacting with friends.^26^

Also, our findings indicate that it would be difficult for these healthcare workers to adopt eHealth and mHealth technologies to help earlier detection and better treatment outcomes. It could also facilitate continuous training and remote support to health workers in rural areas. A systematic review on eHealth and mHealth in developing countries stated that Bangladesh was leading in finding innovative technological ways in their health systems which is now evident in the country having the longest life expectancy and lowest under-five mortality rates.^27, 28^ For a country to easily implement eHealth in primary healthcare facilities, there should be well-designed education and training for all stakeholders including the healthcare providers.^29^ The government also has a role in ensuring access to computers and necessary technical support in these centers. The higher proportion of participants with access to social media compounded with an increase in cell phone and internet technologies use shows that there may be hope for developing countries such as Ghana in incorporating eHealth into their health systems.^30^

The strength of our study is that we enrolled participants from different health centers and data was collected at the participant’s convenient time in their respective workstations. The study also suffered from few limitations. Our study may have suffered response bias because the data was self-reported. The study did not assess the attitude of workers, which could have influenced computer skills and frequency of use. The small sample size of our study is another limitation hence future studies should be at a larger level to have a larger sample to help with generalizability. We also did not find out what social media was used for, future studies should include a qualitative aspect to obtain an in-depth understanding of the attitude and purpose of using social media. Also, more social media platforms including Twitter and Instagram should be explored in future studies.

## Conclusions

The study aimed to assess health workers’ basic computer skills and use of social media and identify specific deficiencies that require training to help them effectively function as above. The results from our study suggest that there is no association between the health profession, years working at a facility, and computer skills or frequency of using computer programs. The findings of the study suggest that healthcare workers’ computer skills and knowledge are relatively low. Health professionals in the health centers need to be trained on basic computer skills and information technology should be incorporated into the health systems for efficient health delivery. Improving computer skills and frequency of using computer programs will help healthcare workers in rural areas to efficiently monitor and assess child health indicators.

## Data Availability

All data produced in the present study are available upon reasonable request to the authors

Supplementary

